# TEST-RETEST RELIABILITY OF MOTOR UNIT FIRING BEHAVIOR DURING MAXIMAL GRIP STRENGTH ASSESSMENTS AMONG OLDER ADULTS

**DOI:** 10.1101/2025.10.12.25337781

**Authors:** Jonathan P. Beausejour, Myles Henderson, Alexander Acevedo, Nathan Bettini, Vanessa Cabrera, Kevan S. Knowles, Meredith Chaput, Grant E. Norte, Joshua C. Kline, Matt S. Stock

## Abstract

Recent evidence suggests that grip strength is a critical biomarker of brain, muscle, and metabolic aging. Presently, neuromuscular properties related to individual motor unit firing properties have not been explored in the context of grip strength assessments in older adults. The purpose of this investigation was to examine the intra- and intersession reliability of linear and exponential regression model coefficients derived from motor unit (MU) peak firing rates (PFRs) and action potential amplitudes (MUAPs) during maximal grip tests among older adults. Thirty-four participants (mean ± SD age = 73 ± 6 years) visited the laboratory on two occasions following a familiarization visit. Two separate maximal grip tests were conducted on participants’ dominant hand during the first visit, and participants revisited the laboratory on a separate day to complete another maximum grip strength test. During each grip test, electromyographical (EMG) signals were recorded from the flexor digitorum superficialis (FDS) muscle and decomposed into individual MUAP trains. Test-retest reliability for linear and exponential regression variables derived from MU PFRs and MUAPs was assessed using intraclass correlation coefficients (ICC_[2,1]_) and paired samples t-test, and validity was determined via calculation of the standard error of the estimate. Y-intercepts (ICC = 0.827) and slopes (ICC = 0.952) from linear models and the decay factor (ICC = 0.866) from the exponential models displayed moderate-to-excellent and good-to-excellent intrasession reliability, respectfully, and poor-to-good intersession reliability (ICCs = 0.646, 0.725, and 0.598, respectively). Paired samples t-tests indicated no group differences in all intra-and inter-session analyses (p ≥ 0.073).

**NEW & NOTEWORTHY:** Reliability of motor unit firing behavior is presented in the context of clinical grip strength assessments among older adults. Despite historical use of exponential modelling to report MUAP-functioned MU FRs, test-retest reliability of linear regression variables were observed to be more reliable and may be a viable analytic approach to characterizing peak MU firing properties between participants and intervention groups when these methodological procedures are adopted.

## INTRODUCTION

Recent evidence suggests that maximal hand grip strength is a critical biomarker of healthy aging.^1^ In aging populations, higher grip strength is strongly associated with overall body strength, bone mineral density, malnutrition, fractures, falls, cognitive impairments, multimorbidity, and quality of life.^2^ Furthermore, grip strength has been shown to predict future changes in physical function.^1^ A 25 year-long longitudinal study by Rantanen et al.^3^ reported that individuals with the lowest baseline grip strength were significantly more likely to exhibit physical limitations over the age of 65.^4^ Due to its relatively simple administration and reliable measurements, grip strength assessments have become commonplace in aging research studies and functional wellness screenings.^5,6^ Consequently, the analysis of grip strength among older adults has become a popular topic of scientific inquisition, with recent investigations highlighting the importance of maintaining skeletal muscle capacity with age to combat functional disabilities.^7,8^

Advancements in surface electromyographical (sEMG) signal processing during skeletal muscle activation have allowed for more robust interpretation of the neural mechanisms responsible for neuromuscular control and force production.^9–11^ With this, important insights can be derived through the assessment of individual MU characteristics recorded during muscle activation. MU characteristics such as recruitment and decruitment, action potential amplitude (MUAP), and FR profiles have been used to investigate adaptions following resistance training protocols^12^, muscular fatigue ^13–16^, and neuromuscular properties between sex and age groups.^17–19^ Typically, trapezoidal, triangular, or patterned isometric muscle contractions are performed to a particular target force intensity (e.g. 80% of maximal voluntary force) during sEMG signal acquisition to quantify MU recruitment threshold, or % of muscle force in which a MU is activated.^9^ MU FRs and action potential amplitudes are then expressed as a function of their respective recruitment threshold, as this allows for more appropriate interpretation of MU properties (e.g. MU size and discharge rate) relative to the active MU pool.^11,20^ Expressing MU properties relative to recruitment threshold have shown to yield reliable test-retest outcomes, thus justifying its approach to reflect changes in MU characteristics following interventional periods and cross sectional analyses across various muscles comparisons across various.^21^ For reference, Colquhoun et al.^21^ reported good-to-excellent inter- and intra-session reliability in the mean firing rates (MFR) versus recruitment threshold (RT) linear relationship recorded in the vastus lateralis,^20–24^ while Parra et al.^22^ demonstrated comparable reliability metrics from EMG signals recorded in the first dorsal interosseous muscle.

Indeed, the MU MFR versus RT and MUAP relationships demonstrate strong correlations (r > .70), and regressed relationships that fall below r < .60 are usually not considered for further statistical analysis.^9,25,26^ MU FRs are inversely related to their recruitment thresholds and action potential amplitudes, respectively, which implicates that smaller MUs recruited earlier during sustained muscle contractions have relatively higher FRs than larger MUs.^20,26,27^ Investigators typically report the linear correlation coefficients (e.g. y intercept, slope, r^2^) of these MFR/MUAP versus RT relationships to interpret neuromuscular characteristics related to neural drive and muscle-specific motor control schemes. As such, it has been proposed that the MFR versus MUAP relationship, specifically, may be best fitted and reported with exponential or power models and their respective coefficients (e.g. scale factors and decay constants related to MU FRs).^25^ Accordingly, the MFR versus MUAP relationship may provide important insights regarding neural drive and other MU-related properties, as its affiliated coefficients appear to be sensitive to assess changes in muscle fiber sizes and ultrasound derived cross-sectional area measures following strength/hypertrophy training protocols and periods of immobilization.^25,28–30^ While recent investigations have also used the MFR versus MUAP relationships to explore MU properties in specific muscle activation strategies^25^ and following orthopedic injury^31^, it is important to note that such investigations employed a cross-sectional design to report MU outcomes. This is noteworthy, as reliability metrics for the MU MFR versus MUAP relationship coefficients have not been formally addressed in the academic record.

Among the challenges in acquiring accurate and appreciable yields in sEMG derived MUAP trains are the alternation and superimposition moments of MUAPs during EMG recordings.^32^ To address this concern, particular contraction types and intensities were recommended to ensure the quality of MU yields during EMG decomposition^9,32^. Specifically, submaximal/maximal isometric contractions completed in controlled activation speeds were employed (e.g. trapezoidal isometric contraction to 50% MVC with 10% MVC/second ramp up and ramp down), with a steady force tracing occurring at the target intensity. Through these conditions, variables such as MU recruitment and de-recruitment thresholds, MUAP, and MU firing properties from the steady portion of the contraction can be reliably extracted, so long as the implemented force pattern was adequately traced by participating research subjects.^9^ As MU FRs usually reported during the steady portion of force, differences in muscle activation/contraction strategies are well documented to influence EMG parameters related to neural drive.^33–36^ As such, evaluations of MU properties in the context of uncontrolled, high velocity maximal contractions offered analytical considerations to report MU firing behavior through their sEMG recorded MUAPs. In comparing MU firings between explosive and controlled maximum contractions in elbow flexors and extensors, Reece et al. (2021) reported strong MFR vs. MUAP power-regressed relationships (r >.65), large MUAP ranges among both muscles, and larger MU yields from explosive contractions decomposed via the Precision Decomposition III algorithm^36^. It is important to note that the authors reported the individual MU mean FRs from a 0.5 epoch within the first instance of a steady torque phase, and MUAP was averaged from separate bipolar sEMG recordings derived from a five-pin sensor array.^36^

There are no published investigations that sought to investigate test-retest reliability of regression coefficients derived from the MU FR versus MUAP during powerful, maximal voluntary isometric contractions (MVIC). Given the established, strong correlations between maximal grip strength performance and overall brain and muscle functionality among older adults, reliably characterizing MU strategies during grip strength assessments is of great interest, especially given the potential of recorded MU parameters to be used as predictive indicators to sarcopenia, or the progressive loss of muscle mass and strength with age^37^. Furthermore, MU properties of implicated hand grip muscles (e.g. flexor digitorum superficialis) during maximal grip force assessments have not been explored, and the sensitive nature of these MU variables in these muscles are unclear related to other muscles commonly reported in the MU literature (e.g. vastus lateralis, first dorsal interosseus, tibialis anterior, ect.).

Considering these important methodological differences, the purpose of this study is to assess the test-retest reliability of MU properties employed during maximum grip strength assessments. With the intention to keep the external validity of the grip force assessment in place, this investigation aims to establish intra- and intersession reliability metrics of regression model coefficients of individual motor unit peak firing rates (PFRs) versus action potential (MUAPs) relationships, specifically^35^. As MU FR versus MUAP relationships are usually reported via exponential and/or power models, the current investigation will also report coefficients from linear regressed relationship to compare differences in model characteristics and derived interpretations. Based on the reported strong correlation between MU FRs and MUAPs, we hypothesize that the slopes, y-intercepts, constant values, and decay factors derived from the PFR versus MUAP relationship will demonstrate moderate-to-excellent intra-session reliability and moderate-to-good intersession reliability.

## MATERIALS AND METHODS

### Experimental Design

This study utilized a repeated measures design to assess the intra-session and inter-session reliability of participants’ maximal grip force and MU PFR versus MUAP relationships. Participants visited the laboratory space on three separate occasions: one familiarization and two testing sessions. During the familiarization visit, participants completed a detailed informed consent document and a comprehensive medical history packet which included the SARC-F questionnaire, Mini-Mental-State Examination-3, and the PAR-Q+ survey. Following completion of study paperwork, participants completed a maximum grip strength assessment, with extra time allocated to ensure adequate practice and proper hand fitting of the hand dynamometer for subsequent testing sessions. Intra-and inter-session reliability was computed for data acquired during the subsequent testing sessions. During the first testing session, two maximum grip strength assessments were administered following a graded warm-up period, with at least ten minutes given between assessments to ensure full recovery. Participants then came back to the laboratory space on another day to complete a third maximal grip assessment. Data collection for each participant was completed at the same time of day (± 1 hour) and lab visits were separated by at least 48 hours, but no longer than two weeks. All participants were asked to refrain from moderate-to-strenuous exercise and alcohol consumption for at least 24 hours, and caffeine consumption at least 4 hours, prior to each laboratory visit.

### Participants

An a priori power analysis was conducted using G*power version 3.1.9.7^38^ for sample size justification. Using a significance criterion of α = 0.05, power = 0.80, and a moderate estimation of correlation among the repeated measures (0.5), the minimum sample size required for this study was computed to be 34 participants. This sample size corresponds with the previous investigations of Colquhoun et al. (2018)^21^ (N = 28) and Parra et al. (2021)^22^ (N = 21), in which intra-session and inter-session reliability of MU data was reported for the vastus lateralis and first dorsal interosseus muscles, respectively. With this, 34 older men (n=17) and women (n=17) were recruited from the neighboring university communities and greater Orlando, FL area (mean ± SD age = 73 ± 6 years, mass = 77.17 ± 15.94 kilograms, height = 168.65 ± 8.34 centimeters, body mass index [BMI] = 26.91 ± 4.00, 28 right-handed). Prior to study enrollment, a comprehensive preliminary health screening was conducted to determine study eligibility. Older adults displaying moderate or severe cognitive impairment (as indicated by the Mini-Mental State Examination) ^39^, or those with a recent history of cancer (< 1 year), any known neurodegenerative (e.g., Parkinson’s, Multiple Sclerosis, Amyotrophic Lateral Sclerosis, etc.) and/or uncontrolled metabolic (e.g., diabetes, hypertension, etc.) disease(s) were excluded. Prospective participants with any history of stroke, or myocardial infarction within the past year, were also excluded. Other exclusionary criteria included: body mass index > 35.0 kg/m^2^, major surgery or hospitalization within the last 6 months, uncontrolled blood clotting disorders, changes in medications or therapies that may alter hormone levels within the previous six months, and history of upper extremity orthopedic surgery or fracture within the previous 6 months. All participants read, understand, and sign informed consent forms detailing study procedures and risks prior to participation, and the procedures were approved by the University of Central Florida’s Institutional Review Board (#7151).

### Height and Body Mass

Standing height and body mass was measured with a stadiometer and physician’s scale, respectively, during the familiarization visit. Measurements were taken while participants stood erect with their heels together and bare foot. Body mass index was computed using participants’ measured height and body mass.

### Maximum Grip Force Assessment

For each visit, maximal grip force assessments were performed on participants’ dominant hand using the Trigno^TM^ Jamar Hand-Grip Dynamometer (Delsys, Inc., Natick, MA, USA). Participants were seated with a neutral position in a customized laboratory chair facing the testing monitor and their hips and back firmly against the chair. Participants’ feet were flat on either the floor, or a level platform, and their elbows were positioned at 90 degrees and supported by an adjustable armrest. The hand dynamometer was placed on participants’ dominant hand and adjusted so the palm side of the dynamometer is secured on the participants’ palm. The front attachment of the dynamometer was adjusted so that it lined with the proximal interphalangeal joints of participants’ hand digits 2-5. Prior to the first maximal grip force assessment of the visit, participants performed three warm-up contractions at perceived submaximal intensities of 50%, 75%, and 90%, respectfully. Following a brief rest period, participants was instructed to squeeze the dynamometer “…as hard and as fast as possible” for five seconds upon a “GO” command from a member of the research team. For each maximal grip force assessment, participants completed three maximal hand grip trails in succession, with a 30-second rest-period between trails. The importance of reacting quickly was emphasized, and strong verbal encouragement was provided throughout testing. Participants were provided visual feedback of their performance via a numerical value (in kilograms) denoting real-time force output during all maximal and submaximal grip contractions. Maximal grip force performance was recorded in newtons (N) and the peak force was determined to the highest 1-second epoch during the duration of each contraction. Hand grip force was sampled at ∼1hz. The highest peak force of the three grip trials performed in each maximal grip assessment was utilized for further analysis. For the familiarization visit, at least one maximum grip force assessment was administered. For the first testing visit, two maximal grip force assessments were assessed, and at least ten minutes of rest was administered to ensure performance recovery from the initial maximal grip force assessment. Only one grip strength assessment was administered during the second testing visit.

### Surface EMG Measurements

Bipolar surface EMG signals were recorded from participants’ dominant flexor digitorum superficialis muscle (FDS), and signals were acquired via a four pin, surface array EMG sensor (Trigno Galileo Sensor, Delsys, Inc., Natick, MA, USA). Before placing the EMG sensor, the location of the FDS sensor placement was identified as 2/3 the distance between the base of the hand to the medial epicondyle of the humerus and marked with a permanent marker^40^. Participants were instructed to flex the proximal interphalangeal joints of their 2^nd^ through 5^th^ digits, and movement of the FDS muscle belly was visually and manually confirmed prior to marker placement. Prior to sensor placement, the skin was shaved with a disposable double-bladed razor, and tape/rubbing alcohol was used to remove dead skin cells, skin oils, and to cleanse the site. The EMG signals from each pin in the array was differentially amplified and filtered with a bandwidth of 20 Hz to 450 Hz, and sampled at 2,222 Hz. Prior to testing, participants performed several submaximal contractions to ensure low baseline noise (≤ 20 mV). EMG signal quality was visually monitored throughout the study, and additional skin preparation or repositioning of the sensors was performed, as necessary.

### Surface EMG Signal Decomposition

Following data acquisition, the four separate filtered surface EMG signals from the muscles was decomposed into their constituent MU action potential trains using the Precision Decomposition III (PD III) algorithm described by De Luca et al. (2006) and further expanded upon by Nawab et al. (2010).^9,11^ Once decomposed, the reconstruct-and-test procedure was used to determine the accuracy of each MU. MUs identified at the >90% accuracy threshold level was included for further analysis.

For each identified MU, two parameters (MU Peak Action Potential Amplitude [mV] and peak firing rate [pps]) was extracted at the one-second epoch where the peak grip force instance occurred. Each MU’s time-varying peak firing rate was computed by passing each train of firings though a one second Hanning window. Linear and exponential regressions were applied on a contraction-by-contraction basis to calculate the slopes/decay factors (pps/ mV), and y-intercepts/constant values (pps) for the PD III derived relationships. Linear and exponential regressions were only applied to include MUs at or above the >90% accuracy threshold level.

### Statistical Analysis

Intra- and inter-session reliability for maximal grip force and the slopes/y-intercepts of the MU peak firing rates (PFR) versus MUAP relationship was quantified with a 2-way random-effects intraclass correlation coefficient model (ICC). Separate ICCs models were used to assess reliability metrics among the number of motor units used for regression modelling, grip force in absolute units (e.g. Newtons), slopes and y-intercepts of the linear regression models, and constant values and decay factors of the exponential models. The 2-way random-effects ICC model was used, specifically, to report generalizable reliability metrics across research centers.^41^ ICCs will be evaluated based on the 95% confident intervals of the ICC point estimation, in which ICC values < 0.50 indicate “poor” reliability, ICCs between 0.50 - 0.75 are indicate “moderate” reliability, ICCs between 0.75 - 0.90 indicate “good” reliability, and ICCs > 0.90 indicate excellent reliability.^42^ The mean square error was used to determine the Standard Error of the Measurement ([SEM] in absolute units and percentage of the grand mean), and the minimal difference value needed to be considered real (MD). As such, SEM and MD values will be calculated based on the formulas outlined in the Weir (2005) report.^41^ In addition, a paired samples t-test (2-tailed) was utilized to examine mean differences in the slopes and y-intercepts within (intra-session) and across (inter-session) testing visits. Prior to statistical analysis, the Shapiro-Wilks and Levene’s tests was used to verify assumptions of normality and homogeneity of variances, respectively. If the data was non-parametric, the paired samples Wilcoxon tests were consulted. Effect size analyses included Cohen’s d statistics to examine pairwise comparisons. Prior to all analyses, the presence of statistical outliers was objectively checked by using the absolute deviations of ± 2.5 around the median.^43^ Small, medium and large effect sizes corresponded to 0.20, 0.50, and 0.80, respectively.^44^ Finally, chi-square tests of independence was conducted to examine differences in the contraction times where peak grip force was achieved during each grip assessment. A customized reliability excel spreadsheet and JASP statistical software (JASP 19.3, University of Amsterdam, Amsterdam, NL) was used to conduct all ICC point estimates and paired samples t-tests.

## RESULTS

### Motor Unit Decomposition Characteristics

A total of 2426 MUs was identified by the PD III algorithm, and 867 MUs were removed when the 90% accuracy threshold was applied. For this analysis, a total of 1578 MUs were included, with an average of 15.5 ± 5.0 MUs observed for each contraction (range: 28 to 6). From the 68 contractions analyzed in the study, two contractions utilized an accuracy threshold of 80% due to the insufficient number of motor units available at the 90% accuracy threshold (11 MUs were used for each of these contractions, respectfully). Average coefficients of determination (R^2^) for linear and exponential regression models were observed to be 0.97 ± 0.02 (range: 0.99 to 0.86) and 0.94 ± 0.03 (range: 0.98 to 0.79), respectively. **Table 1** presents descriptive data for the key linear and exponential variables observed in this study.

**Table 1.** Descriptive data for key linear and exponential regression model variables across contractions.

|  | Number of Motor Units | Grip Force (N) | Y-Intercept (pps) | Slope (pps/mV) | Constant (pps) | Decay Factor (pps/mV) |
| --- | --- | --- | --- | --- | --- | --- |
| **Visit 1:**<br>**Contraction 1** | 14.82 ± 4.54 | 321.70 ± 103.38 | 28.63 ± 2.43 | -53834.41<br>25189.22 | 32.91 ± 4.92 | -3715.94<br>± 2254.87 |
| **Visit 1:**<br>**Contraction 2** | 15.18 ± 5.36 | 318.51 ± 104.06 | 28.45 ± 2.52 | -56160.91 ±<br>23415.47 | 32.36 ± 6.32 | -3960.76<br>± 2208.79 |
| **Visit 2:**<br>**Contraction 1** | 16.412 ± 5.11 | 325.81 ± 97.38 | 28.79 ± 2.86 | -51468.35 ±<br>20571.87 | 32.55 ± 3.92 | -3485.84<br>± 1676.15 |

Mean ± standard deviations observed for the number of motor units used to create the linear and exponential regression models, grip force expressed in newtons (N), y-intercept values derived from the linear regression models expressed as motor units' peak pulses per second (pps), slope values derived from the linear regression models expressed in (pps) functioned peak motor unit action potential amplitudes (pps/mV), constant values derived from the exponential regression models expressed as motor units' peak pulses per second (pps) and decay factors derived from the exponential regression models expressed in (pps) functioned peak motor unit action potential amplitudes (pps/mV).

### Intrasession Reliability

**Table 2** presents the intrasession reliability statistics for the number of motor units used for the regression models, grip force, y-intercept, slope, constant value, and decay factor variables. The chi-square test examining contraction times where maximal grip force was achieved indicated no differences between the grouped measurement instances (*χ*^2^= 1.588, *p =* 0.811)

**Table 2.**
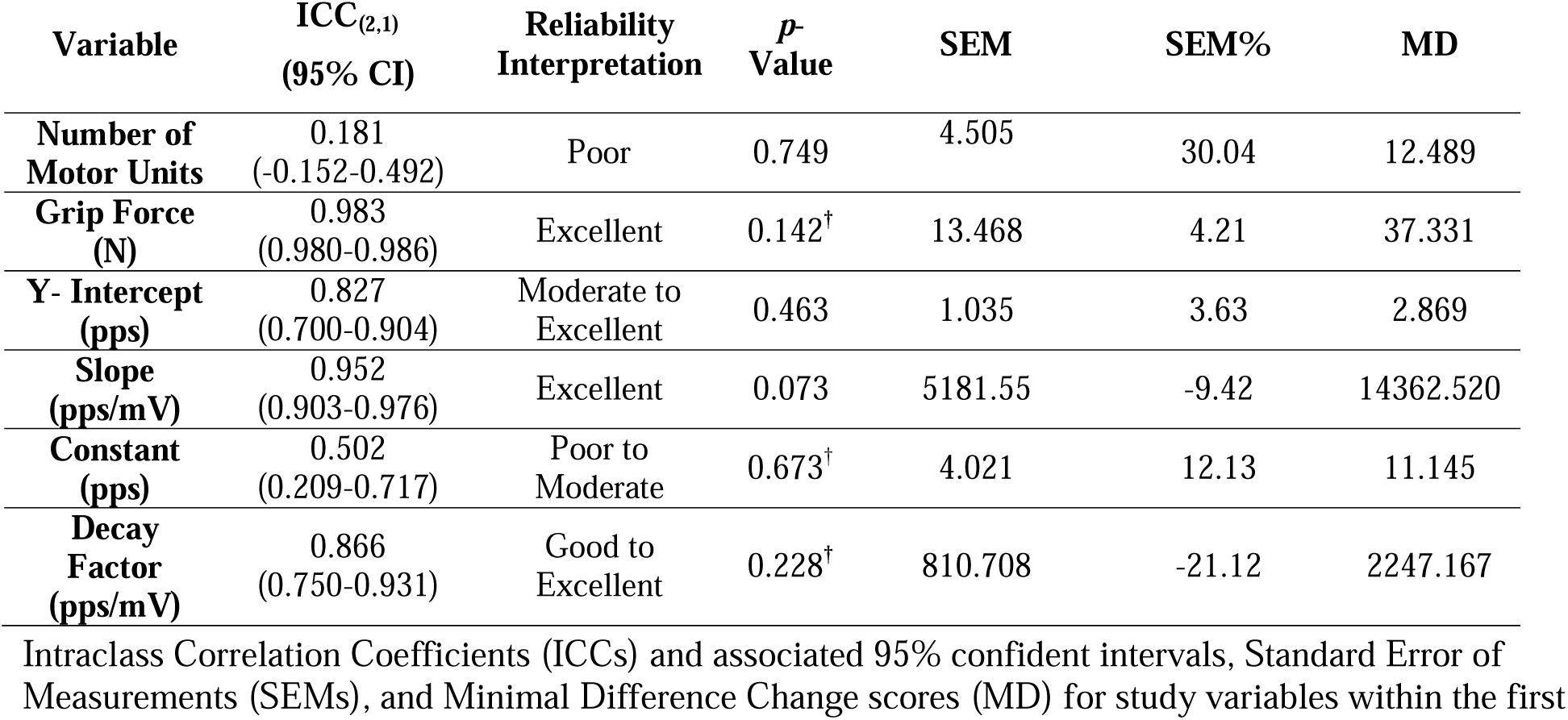
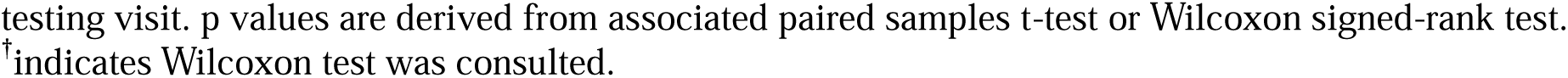
Intraclass correlation coefficients and paired samples test results for study variables (Intrasession)

| Variable | ICC<sub>(2,1)</sub><br>(95% CI) | Reliability Interpretation | p-Value | SEM | SEM% | MD |
| --- | --- | --- | --- | --- | --- | --- |
| **Number of Motor Units** | 0.181<br>(-0.152-0.492) | Poor | 0.749 | 4.505 | 30.04 | 12.489 |
| **Grip Force (N)** | 0.983<br>(0.980-0.986) | Excellent | 0.142<sup>†</sup> | 13.468 | 4.21 | 37.331 |
| **Y- Intercept (pps)** | 0.827<br>(0.700-0.904) | Moderate to Excellent | 0.463 | 1.035 | 3.63 | 2.869 |
| **Slope (pps/mV)** | 0.952<br>(0.903-0.976) | Excellent | 0.073 | 5181.55 | -9.42 | 14362.520 |
| **Constant (pps)** | 0.502<br>(0.209-0.717) | Poor to Moderate | 0.673<sup>†</sup> | 4.021 | 12.13 | 11.145 |
| **Decay Factor (pps/mV)** | 0.866<br>(0.750-0.931) | Good to Excellent | 0.228<sup>†</sup> | 810.708 | -21.12 | 2247.167 |

Intraclass Correlation Coefficients (ICCs) and associated 95% confident intervals, Standard Error of Measurements (SEMs), and Minimal Difference Change scores (MD) for study variables within the first
testing visit. p values are derived from associated paired samples t-test or Wilcoxon signed-rank test. <sup>†</sup> indicates Wilcoxon test was consulted.

### Number of Motor Units (Intrasession)

The number of motor units displayed poor intrasession reliability (ICC = 0.181 [-0.152 – 0.492]) and the corresponding paired samples t-test indicated no significant differences between grouped measurement instances (*p =* 0.749, mean difference = -0.35; *d =* -0.055), as shown in Figure 2A.

**Figure 1.**
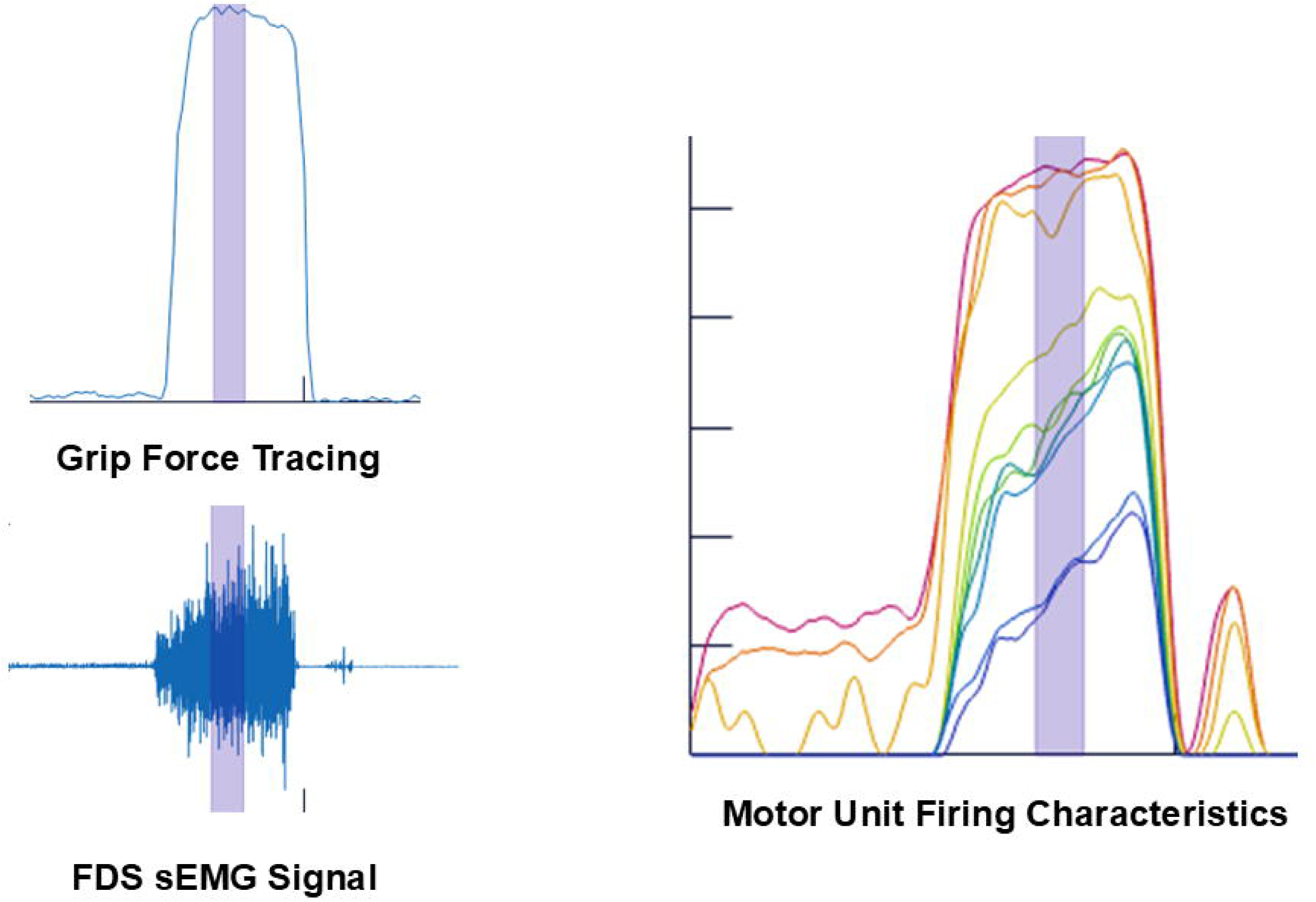
sEMG data extraction: MU data extraction for grip assessment corresponded to the 1-second epoch (shaded area) where the highest peak grip force was achieved.

**Figure 2.**
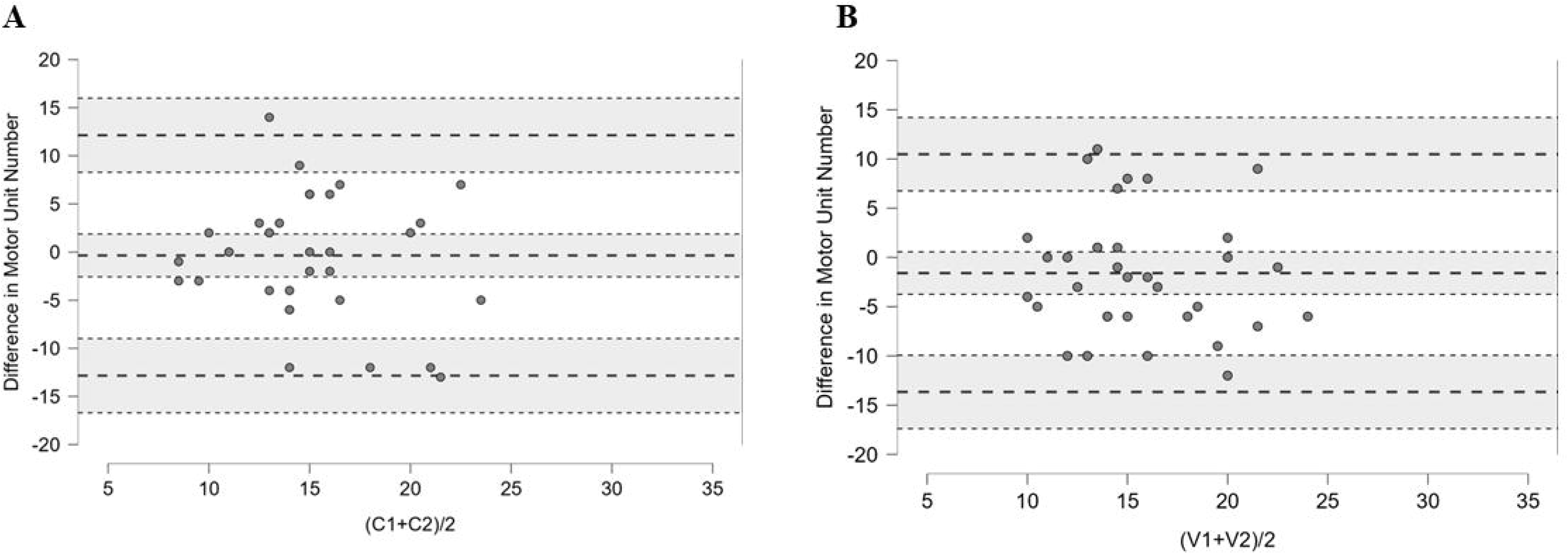
Bland-Altman Plots of Number of Motor Units: JASP Bland Altman plot displays agreement characteristics in the number of motor units used to create linear and exponential models between contractions acquired within (A) and between (B) visits for the 34 participants. The x-axis represents the mean number of motor units available across both contractions during testing visit 1(C1 and C2) and between visits (V1 and V2). The y-axis represents the mean differences of the number of motor units observed in each participant (dots). For both (A) and (B), the middle-dashed line represents the overall group bias, and the top/bottom dashed lines represent the 95% limits of agreement (LOA). (A) Bias [LOA] = -0.35; [-12.84 – 12.14], (B) = -1.59 [-13.67 – 10.45]. Shaded areas represent the 95% CI of the bias and limits of agreement.

### Maximal Grip Force (Intrasession)

Participants’ maximal grip force performance displayed excellent intrasession reliability (ICC = 0.983 [0.980 – 0.986]) and the corresponding Wilcoxon signed-rank test indicated no significant differences between grouped measurement instances (*p =* 0.142, mean difference = - 3.19; *d =* 0.299), as shown in Figure 3A.

**Figure 3.**
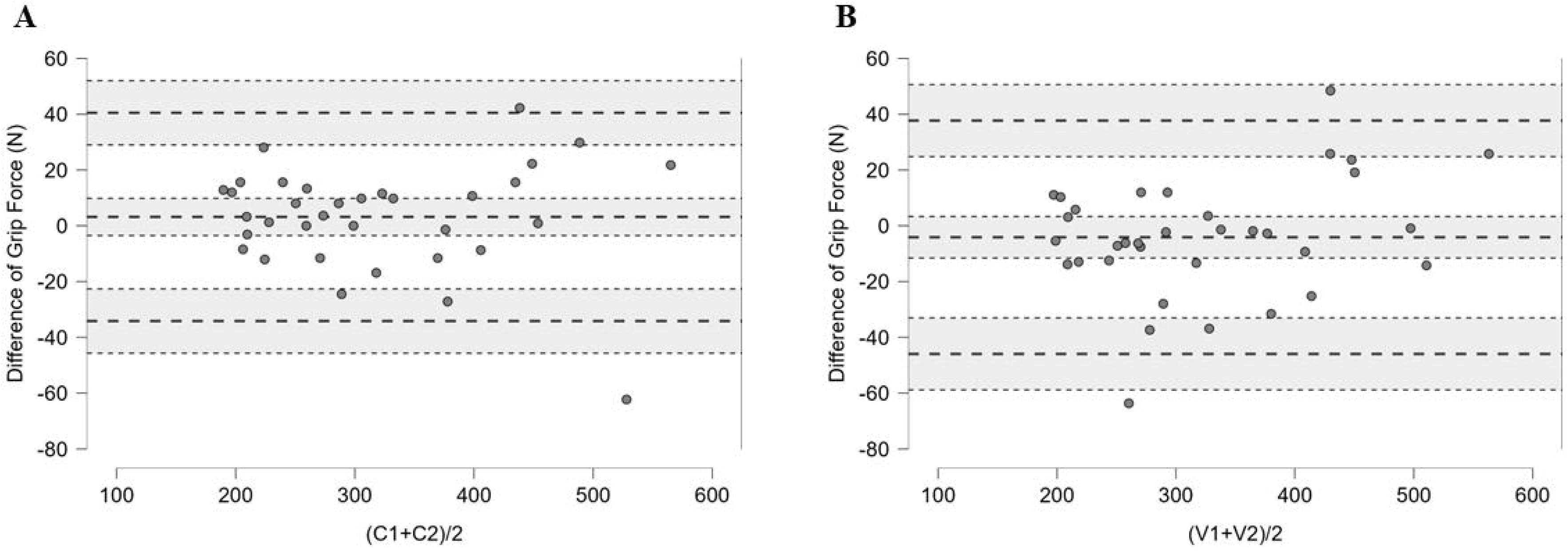
Bland-Altman Plots of Grip Force (Newtons): JASP Bland Altman plot displays agreement characteristics in grip force acquired within (A) and between (B) visits for the 34 participants. The x-axis represents the participants’ mean grip force across both contractions during testing visit 1 (C1 and C2) and between visits (V1 and V2). The y-axis represents the mean differences of grip force observed in each participant (dots). For both (A) and (B), the middle-dashed line represents the overall group bias, and the top/bottom dashed lines represent the 95% limits of agreement (LOA). (A) Bias [LOA] = 3.19; [-34.14 – 40.52], (B) = -4.11 [-45.94 – 37.72]. Shaded areas represent the 95% CI of the bias and limits of agreement.

### Linear Model: Y-Intercept (Intrasession)

Y-intercepts derived from the linear regression models displayed good-to-excellent intrasession reliability (ICC = 0.827 [0.700 – 0.904]) and the corresponding paired samples t-test indicated no significant differences between grouped measurement instances (*p =* 0.463, mean difference = -0.19; *d =* 0.127) as shown in Figure 4A.

**Figure 4.**
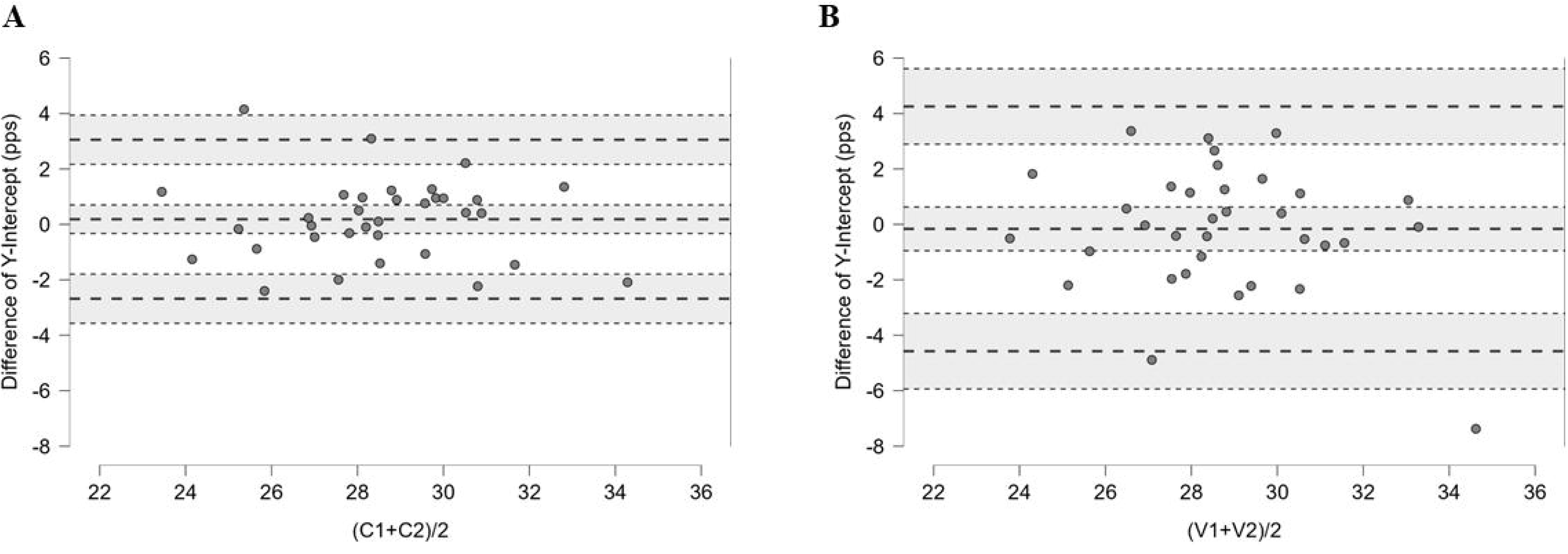
Bland-Altman Plots of Y-Intercept: JASP Bland Altman plot displaying agreement characteristics in y-intercepts observed from MU PFR vs. MUAP linear regressions acquired within (A) and between (B) visits for the 34 participants. The x-axis represents the participants’ mean y-intercepts across both contractions during testing visit 1 (C1 and C2) and between visits (V1 and V2). The y-axis represents the mean differences of y-intercepts observed in each participant (dots). For both (A) and (B), the middle-dashed line represents the overall group bias, and the top/bottom dashed lines represent the 95% limits of agreement (LOA). (A) Bias [LOA] = 0.19; [-2.68 – 3.06], (B) = -0.16 [-4.58 – 4.26]. Shaded areas represent the 95% CI of the bias and limits of agreement.

### Linear Model: Slope (Intrasession)

Slopes derived from the linear regression models displayed excellent intrasession reliability (ICC = 0.952 [0.903 – 0.976]) and the corresponding paired samples t-test indicated no significant differences between grouped measurement instances (*p =* 0.073, mean difference = - 2326.50; *d =* 0.317) as shown in Figure 5A.

**Figure 5.**
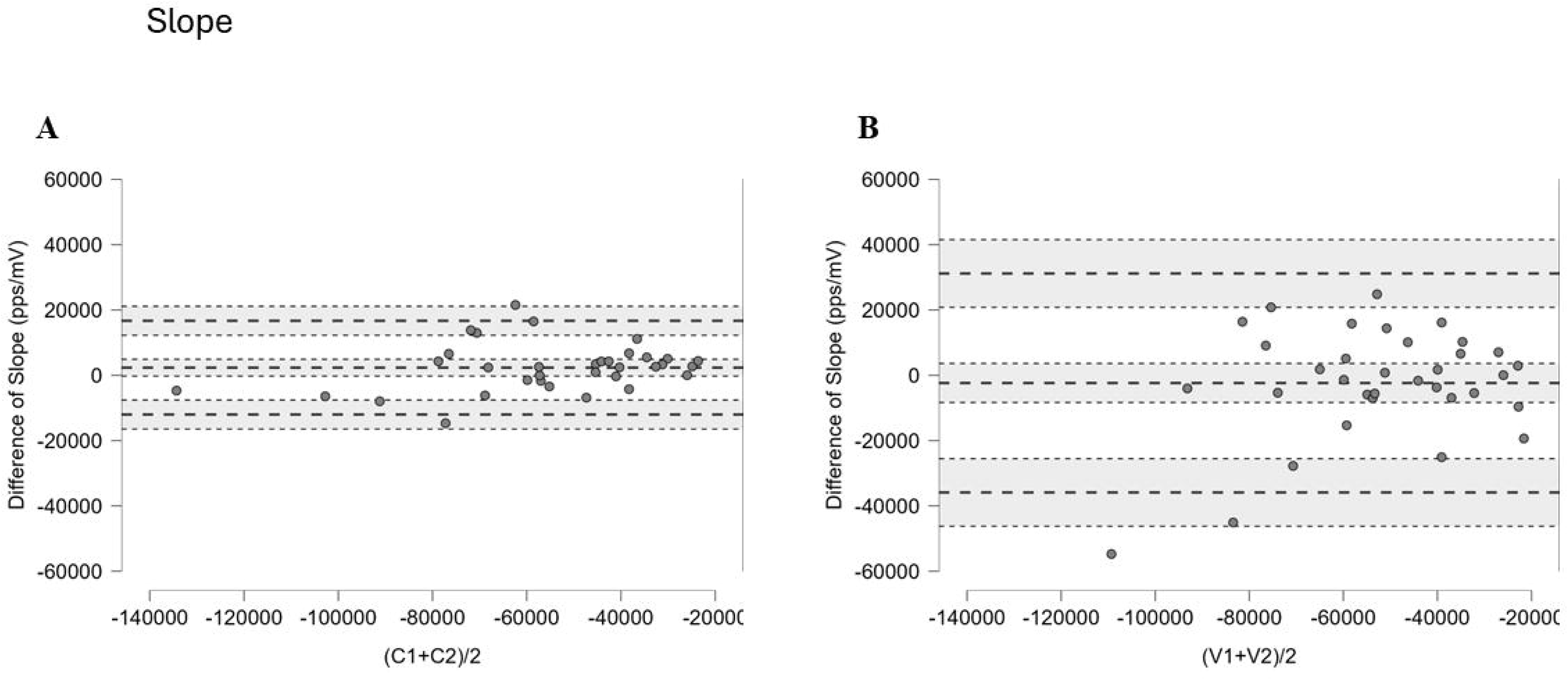
Bland-Altman Plots of Slope: JASP Bland Altman plot displaying agreement characteristics in slopes observed from MU PFR vs. MUAP linear regressions acquired within (A) and between (B) visits for the 34 participants. The x-axis represents the participants’ mean slopes across both contractions during testing visit 1 (C1 and C2) and between visits (V1 and V2). The y-axis represents the mean differences of slopes observed in each participant (dots). For both (A) and (B), the middle-dashed line represents the overall group bias, and the top/bottom dashed lines represent the 95% limits of agreement (LOA). (A) Bias [LOA] = 2326.50; [-12036.02 – 16689.02], (B) = 2366.06 [-35906.83 – 31174.71]. Shaded areas represent the 95% CI of the bias and limits of agreement.

### Exponential Model: Constant Value (Intrasession)

Constant values derived from the exponential regression models displayed poor-to-moderate intrasession reliability (ICC = 0.502 [0.209 – 0.717]) and the corresponding Wilcoxon signed-rank test indicated no significant differences between grouped measurement instances (*p =* 0.673, mean difference = 0.46; *d = -*0.086) as shown in Figure 6A.

**Figure 6.**
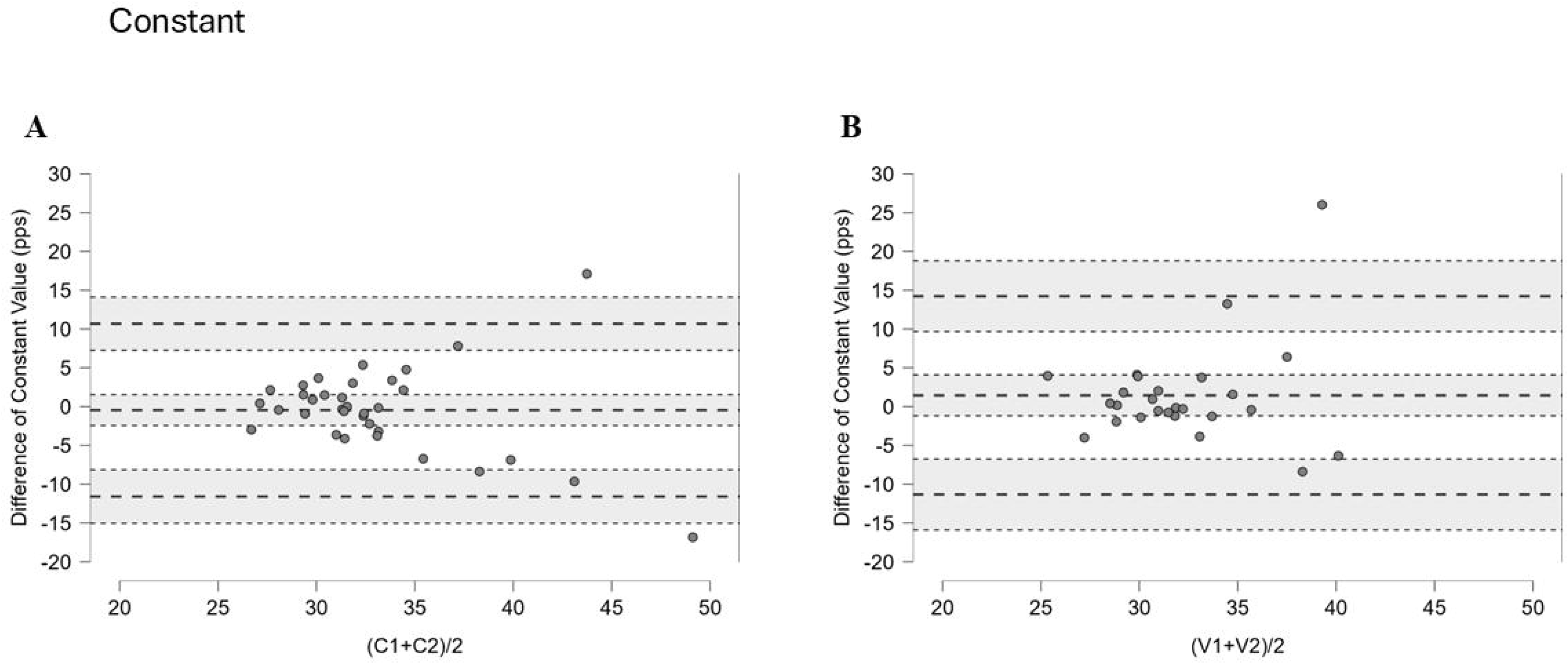
Bland-Altman Plots of Constant Values: JASP Bland Altman plot displaying agreement characteristics in constant values observed from MU PFR vs. MUAP exponential regressions acquired within (A) and between (B) visits for the 34 participants. The x-axis represents the participants’ mean constants across both contractions during testing visit 1 (C1 and C2) and between visits (V1 and V2). The y-axis represents the mean differences of constants observed in each participant (dots). For both (A) and (B), the middle-dashed line represents the overall group bias, and the top/bottom dashed lines represent the 95% limits of agreement (LOA). (A) Bias [LOA] = -0.46; [-11.60 – 10.69], (B) = 1.447 [-11.33 – 14.23]. Shaded areas represent the 95% CI of the bias and limits of agreement.

### Exponential Model: Decay Factor (Intrasession)

Decay factors derived from the exponential regression models displayed good-to-excellent intrasession reliability (ICC = 0.866 [0.750 – 0.931]) and the corresponding paired samples t-test indicated no significant differences between grouped measurement instances (*p =* 0.228, mean difference = -244.79; *d =* 0.239), as shown in Figure 7A.

**Figure 7.**
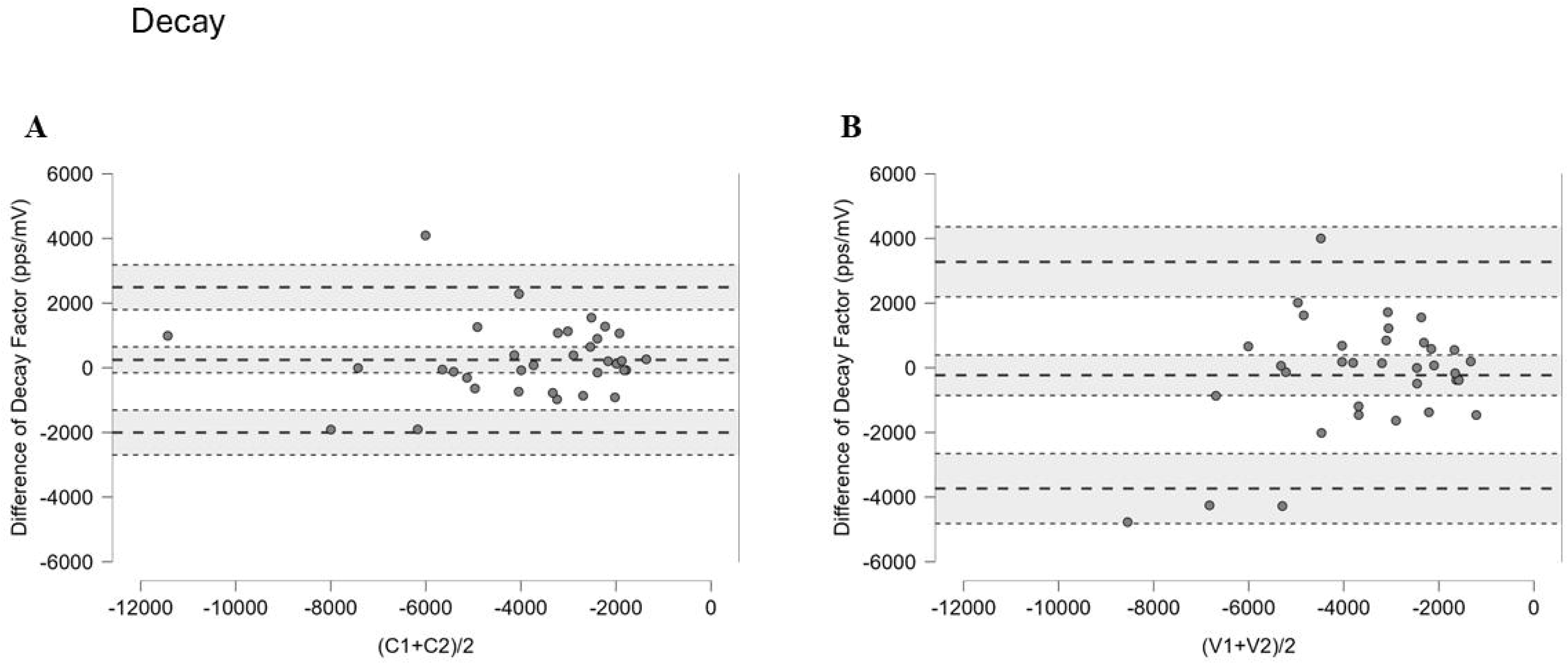
Bland-Altman Plots of Decay Factor: JASP Bland Altman plot displaying agreement in decay factors observed from MU PFR vs. MUAP exponential regressions acquired within (A) and between (B) visits. The x-axis represents the participants’ mean decay factors across both contractions during testing visit 1 (C1 and C2) and between visits (V1 and V2). The y-axis represents the mean differences of decay factors observed in each participant (dots). For both (A) and (B), the middle-dashed line represents the overall group bias, and the top/bottom dashed lines represent the 95% limits of agreement (LOA). (A) Bias [LOA] = 244.79; [-2002.37 – 2491.96], (B) = -230.11 [-3736.83 – 3276.62]. Shaded areas represent the 95% CI of the bias and limits of agreement.

### Intersession Reliability

**Table 3** presents the intersession reliability statistics for the number of motor units used for the regression models, grip force, y-intercept, slope, constant value, and decay factor variables. Mean ± SD days between visits = 5.26 ± 3.35 (range: 2 to 14 days). The chi-square test examining contraction times where maximal grip force was achieved indicated no differences between the grouped measurement instances (χ^2^ = 1.579, *p =* 0.812).

**Table 3.** Intraclass correlation coefficients and paired samples test results for study variables (Intersession)

| Variable | ICC<sub>(2,1)</sub><br>(95% CI) | Reliability<br>Interpretation | p-Value | SEM | SEM% | MD |
| --- | --- | --- | --- | --- | --- | --- |
| Number of Motor Units | 0.183<br>(-0.146-0.481) | Poor | 0.142 | 4.356 | 27.89 | 12.074 |
| Grip Force (N) | 0.977<br>(0.973-0.981) | Excellent | 0.221<sup>†</sup> | 15.091 | 4.66 | 41.831 |
| Y- Intercept (pps) | 0.646<br>(0.429-0.795) | Poor to Good | 0.680 | 1.593 | 5.55 | 4.415 |
| Slope (pps/mV) | 0.725<br>(0.520-0.852) | Moderate to Good | 0.853<sup>†</sup> | 12100.460 | -22.98 | 33540.770 |
| Constant (pps) | -0.115<br>(-0.334-0.334) | Poor | 0.543<sup>†</sup> | 4.688 | 14.32 | 12.994 |
| Decay Factor (pps/mV) | 0.598<br>(0.334-0.777) | Poor to Good | 0.853<sup>†</sup> | 1265.118 | -35.13 | 3506.727 |

Intraclass Correlation Coefficients (ICCs) and associated 95% confident intervals, Standard Error of Measurements (SEMs), and Minimal Difference Change scores (MD) for study variables between visits. p values are derived from associated paired samples t-test or Wilcoxon signed-rank test. <sup>†</sup> indicates Wilcoxon test was consulted.

### Number of Motor Units (Intersession)

The number of motor units displayed poor intersession reliability (ICC = 0.183 [-0.146 – 0.481]) and the corresponding paired samples t-test indicated no significant differences between grouped measurement instances (*p =* 0.142, mean difference = -1.59; *d = -*0.258), as shown in Figure 2B.

### Maximal Grip Force (Intersession)

Participants’ maximal grip force performance displayed excellent intersession reliability (ICC = 0.977 [0.973 – 0.981]) and the corresponding Wilcoxon signed-rank test indicated no significant differences between grouped measurement instances (*p =* 0.221, mean difference = 4.11; *d =* -0.244), as shown in Figure 3B.

### Linear Model: Y-Intercept (Intersession)

Y-intercepts derived from the linear regression models displayed poor-to-good intersession reliability (ICC = 0.646 [0.429 – 0.795]) and the corresponding paired samples t-test indicated no significant differences between grouped measurement instances (*p =* 0.680, mean difference = 0.16; *d =* -0.071) (Figure 4B).

### Linear Model: Slope (Intersession)

Slopes derived from the linear regression models displayed moderate-to-good intersession reliability (ICC = 0.725 [0.520 – 0.852]) and the corresponding Wilcoxon signed-rank test indicated no significant differences between grouped measurement instances (*p =* 0.853, mean difference = 2366.06; *d =* -0.039) (Figure 5B).

### Exponential Model: Constant Value (Intersession)

Constant values derived from the exponential regression models displayed poor intersession reliability (ICC = -0.115 [-0.334 – 0.334]) and the corresponding Wilcoxon signed-rank test indicated no significant differences between grouped measurement instances (*p =* 0.543, mean difference = 0.-36; *d = -*0.123) (Figure 6B).

### Exponential Model: Decay Factor (Intersession)

Decay factors derived from the exponential regression models displayed poor-to-good intrasession reliability (ICC = 0.598 [0.334 – 0.777]) and the corresponding Wilcoxon signed-rank test indicated no significant differences between grouped measurement instances (*p =* 0.853, mean difference = 230.11; *d =* -0.039) (Figure 7B).

## DISCUSSION

This investigation sought to examine test-retest intra- and intersession reliability of linear and exponential model coefficients derived from the MU PFR versus MUAP relationship. In our analysis, Y-intercepts (ICC = 0.827) and slopes (ICC = 0.952) from linear models and the decay factor (ICC = 0.866) from the exponential models displayed good-to-excellent intrasession reliability, and poor-to-good intersession reliability (ICC = 0.646, 0.725, and 0.598, respectively). Constant values from the exponential models displayed poor-to-moderate and poor intra and intersession reliability, respectively. Paired samples t-tests indicated no group differences in all intra-and inter-session analyses (p ≥ 0.073), indicating stable grouped differences within and between testing sessions and substantial within-participant measurement variation. Of note, all reliability metrics were observed despite poor test-retest reliability observed for the number of motor units used to create regression models. Below we discuss our interpretation, noteworthy perspectives, and statistical considerations for future investigations interested in adopting the analytical approach implemented in this study.

As we hypothesized, very strong relationships were observed for both linear and exponential models, however intra- and intersession reliability metrics for key regression variables differed, with lower ICCs observed in the intersession analyses. While the investigations of Colquhoun et al.^21^ and Parra et al.^22^ reported good to excellent intra- and inter-session reliability of recruitment threshold (RT)-functioned MU y-intercepts and slopes (ICCs >.0.766), both studies employed strict MU criteria to assess MU reliability metrics of submaximal contractions. Herein, moderate-to-excellent intrasession reliability markers were observed for key regression coefficients, which may signal the need to use specific MU criteria to improve intersession reliability metrics. Other factors may have contributed to the diminished intersession reliability metrics, such as the muscle chosen to record MUs (e.g. FDS versus vastus lateralis and first dorsal interosseous muscles), the specific type of EMG sensor used, and the inherit variability of EMG signal acquisition between visits (e.g. specific sensor location, sensor pressure, and skin conduction properties). While inherit variation related to EMG acquisition steps exist across all MU investigations, these differences may play a larger role when reporting MU firing rate as a function of their respective motor unit action potential amplitude in older adults.^45^ Regardless of these factors, group measurements of the linear and regression coefficient variables were not different, as indicated by the insignificant paired samples tests. As such, a case may be presented that regression model variables may fall subject, more so, to within-subject variation of measurement outcomes (e.g. grip force variance between contractions) as opposed to systemic EMG acquisition error. It is noteworthy that the present reliability metrics were achieved despite poor reliability in the number of motor units used to create the regression models, although reliability metrics of motor units used to create models are not typically reported in MU reliability investigations.

Historically, MU firing rates reported as a function of their MUAP are expressed via exponential or power regression models, as MUAP indicates the relative size of the MU when RTs are not used^36^. MU RT values are obtained, primarily, through the controlled contraction mechanism typically employed in MU studies. As such, between-group comparisons in MU properties can be investigated by standardizing the %RT across participants. In our investigation, recruitment threshold measures were not obtained due to the deliberate attempt to capture motor unit properties during a powerful muscle contraction. To control how MU properties are standardized across participants, MUs were extracted from the one-second epoch where the highest force was achieved, thus rendering our MU FR characteristics to be reported as a function of their respective MUAPs. Interestingly, y-intercept values derived from the linear regression models demonstrated superior test-retest reliability compared to the constant values, while comparable reliability markers were noted for slope and decay factors. With this, our investigation provides evidence that reporting MUAP-functioned MU PFRs via linear regression models are a viable analytic approach to characterizing MU firing properties between participants and groups.

A surprising observation in the present study was the superior reliability metrics observed for the slope and decay values from the linear and exponential models, respectively, despite poor test-retest reliability for the number of motor units. In the context of MU literature, slope and decay values reflect the nature of MU characteristics across the recorded motor unit pool, allowing for inferences regarding the firing behavior of lower-threshold MUs (e.g. smaller) and higher threshold MUs (e.g. larger). In investigations that include MU RTs, the relative range of low and high threshold motor units are primarily based on the MU’s %RTs observed during a controlled contraction, as explained by the general size principle of muscle activation.^46^ Herein, the size of identified MUs may be inferred by the magnitude of their MUAP, with comparable linear and exponential model characteristics to those that report MU FRs as a function their %RT. As such, the analytical approach implemented in this investigation may be sufficient to reliably track adaptations of specific MUs that influence muscle performance and capacity (e.g. firing characteristics of higher threshold motor units). This is especially relevant given the current novelty of describing MU properties in the context of powerful muscle activations, as opposed to traditionally controlled muscle activations.

An interesting observation among reliability metrics was the lack of significant differences among grouped measurements in all within and between visit comparisons. With ICC interpretations ranging from poor-to-excellent in all study variables, a case may be presented that ICC metrics are largely influenced by within-participant variation between measurements. It is widely recognized that variations in MU properties within and between participants make between group comparisons, or investigations involving an intervention component, difficult to quantify. Given the reliability metrics observed, statistical methods and study designs that account for within-subject variation in motor unit properties may better address the inherent variability of neurophysiological data than approaches relying solely on group mean comparisons.

This study has multiple considerations that should be considered. First, peak firing rates and motor unit action potentials were extracted as primary MU characteristics. As peak firing rates have been proposed to indirectly implicate rate of force development markers^35^, the rationale to use peak values was rooted in the nature of the grip force assessment. As such, MU processing procedures differ from what was previously reported, most notably that of Reece et al.^36^, where the authors reported the mean MU FRs and MUAPs from a 0.5 second epoch within a stable portion of the maximal contraction. Another limitation includes the relatively low sample rate of the hand-grip dynamometer used for this investigation, as this sampling rate is significantly less than sampling rates reported for typical dynamometer-acquired force outcomes. MUAPs used for regression models were not normalized to participant-specific sEMG characteristics to a reference contraction instance, thus reducing the relative reporting of MUAP measures acquired in our analysis. Furthermore, reporting of MU properties used MUs identified with the 90% accuracy threshold in all but 2 contractions, where the 80% accuracy thresholds were used to obtain adequate MU yields. Lastly, we cannot fully guarantee the absence of potential crosstalk of neighboring muscles during sEMG acquisition of the flexor digitorum superficialis muscle and that the same motor units were recorded across the included contractions instances within and between lab visits.

## CONCLUSION

In conclusion, this investigation is among the first studies to establish MU reliability metrics in context-specific, forceful muscle activations, and sought to use a novel methodological approach to reliably tract MU characteristics in a maximal grip force assessment among older adults. Our results demonstrated moderate-to-excellent intrasession reliability in linear regression derived y-intercepts and slopes, as well as good-to-excellent intrasession reliability in exponential regression-derived decay factors. Intersession ICCs were lower across all key regression model variables, and may be mostly attributed to within-subject differences between measurement instances. As such, adopting statistical approaches and associated study designs that properly account for the within-subject variation in MU properties is recommended. Despite the historical use of exponential modelling to report MUAP-functioned MU FRs, test-retest reliability of linear regression variables were observed to be good-to-excellent, and may be considered a viable analytic approach to characterizing peak MU firing properties between participants and intervention groups when these methodological procedures are adopted. Importantly, all reliability metrics were observed despite poor test-retest reliability in the number of motor units used for regression modeling.

## Data Availability

All data produced in the present study are available upon reasonable request to the authors

## ACKNOWLEDGMENTS

The authors extend their sincerest gratitude to all of the research participants that took part in this study, and to Legacy Pointe at UCF Elderly Living Facility for providing space to conduct our research project.

Preprint is available at http://doi.org/xyz

## AUTHOR CONTRIBUTIONS

JPB and MSS conceived and designed the research study. JPB, MH, AA, NB, and VC performed the research experiment and analyzed the subsequent data. JPB, MSS, MC, GEN, JCK interpreted results of the study. JPB drafted initial draft and prepared figures and tables. MSS, MH, AA, NB, VC, KK, MC, GEN, and JCK edited and revised the manuscript and approved the final version of the manuscript.

